# Urinary prostaglandin metabolites as biomarkers for human labour: Insights into future predictors

**DOI:** 10.1101/2024.11.28.24318140

**Authors:** Eilidh M. Wood, Kylie K. Hornaday, Matthew Newton, Melinda Wang, Stephen L. Wood, Donna M. Slater

## Abstract

Prostaglandins and other related molecules in the eicosanoid family have long been implicated in the process of both term and preterm labour. Although, exactly which eicosanoids are involved and whether they have utility as biomarkers for labour, remains to be shown. The objective of this study was to determine whether urinary prostaglandins and related molecules a) change with labour and/or cervical changes, at term and preterm, and/or b) are associated with timing of delivery in individuals with threatened preterm labour. Pregnant individuals were recruited into the following groups: n=32 term non-labour, n=49 term labour, n=15 preterm non-labour controls, n=43 threatened preterm labour with preterm delivery, and n=44 threatened preterm labour with term delivery. Metabolites of prostaglandins PGE_2_, PGF_2α_, PGD_2_, and PGI_2_ as well as 8-isoprostane were measured by ELISA. In addition, 147 eicosanoids were measured in a small (n=24) subset of these samples using a mass-spectrometry based targeted lipidomics panel. At term labour prostaglandin PGF_2α_ and PGE_2_ and PGF_2α_ metabolites were increased compared to term non-labour. There were no changes in any prostaglandin metabolites prior to labour onset. Prostaglandin I_2_ metabolite was lower in individuals with threatened preterm labour who delivered preterm compared to those who went on to deliver at term. In our discovery cohort, we identified 20 additional eicosanoids as highly expressed in maternal urine, include members of the prostaglandin, hydroxyeicosatetraenoic acid (HETE), epoxyeicosatrienoic acid (EET), dihydroxy-octadecenoic acid (DiHOME), dihydroxy-eicosatrienoic acid (diHETrE), isoprostane, and nitro fatty acid eicosanoid families. In conclusion, we did not identify any prostaglandins that would have utility as predictors for term or preterm labour, however, we have identified diverse eicosanoids that have not been previously explored in the context of pregnancy and labour, highlighting novel areas for biomarker research.

## Introduction

Prostaglandins are biological mediators with key roles in reproduction, including pregnancy and parturition. Prostaglandins may contribute to the labour process by promotion of cervical ripening [1–3], uterine smooth muscle contraction [4,5] and membrane rupture [6–8]. In particular, the prostaglandins PGE_2_ and PGF_2α_ and their synthetic analogues are clinically important in obstetrics and have been used to induce and augment labour since the late 1960s [9]. In addition, while not used clinically in obstetrics, PGI_2_ and PGD_2_ are also implicated in mechanisms that may play a role in human uterine relaxation and contraction [4,10–12]. Prostaglandins belong to a larger family of lipid signaling molecules called eicosanoids, and some of these related molecules, including the isoprostanes, which constitute a group of prostaglandin-like compounds, may also be involved in pregnancy and labour [13–16].

The mechanisms that contribute to the onset of human labour are not well understood. This limits our ability to clinically manage labour, most significantly in the case of preterm labour, which is a leading cause of neonatal morbidity and mortality worldwide [17]. Although many biomarkers for preterm labour have been proposed, none so far have sufficient predictive ability for clinical use [18,19]. Importantly, only ∼50% of individuals presenting with symptoms of preterm labour (threatened preterm labour) (TPTL), deliver prematurely [20,21], yet there is a lack of accurate measures for assessing which of these individuals will deliver preterm and which will deliver at term. A sensitive and specific biomarker for preterm delivery in the context of threatened preterm labour would allow for targeted clinical management and allocation of healthcare resources towards those at high risk of delivering preterm.

Available data on prostaglandin levels in maternal biofluids (urine, blood, amniotic fluid) provides some evidence that prostaglandin levels increase with labour [22], suggesting a possible role for prostaglandins as biomarkers for labour. Circulating prostaglandins are rapidly metabolized and secreted into the urine, which presents an opportunity to assess prostaglandin metabolites as potential biomarkers during pregnancy. Further, urine can be easily and non-invasively collected in relatively large volumes for biomarker analysis. We hypothesize that urinary prostaglandin levels are associated with labour and that possible alterations in prostaglandin levels associated with preterm labour are detectable in maternal urine. However, it is unknown whether urinary prostaglandin levels increase prior to labour onset and/or with the onset of true labour. A better understanding of how prostaglandins and/or their metabolites are involved in labour may allow for identification of biomarkers for preterm labour and preterm birth and possibly better targets for prevention and treatment of this condition. In addition, other, non-prostaglandin, eicosanoids may also be involved in the labour process, and a more complete understanding of the range of eicosanoids present in maternal circulation during pregnancy and labour may open new avenues for biomarker discovery.

Therefore, the first aim of this study is to determine whether the urinary prostaglandins PGF_2α_ and prostaglandin metabolites PGFM, PGEM, PGIM, and tetranor-PGDM (t-PDGM), as well as 8-isoprostane, a) change with labour and/or cervical changes, at term and preterm, and b) are associated with timing of delivery in those with TPTL. The second aim of this study is to perform a discovery analysis using a mass-spectrometry-based eicosanoid panel on a subset of samples and determine if any additional eicosanoids are associated with labour.

## Methods

### Participants and sample collection

Institutional ethics was obtained for the study (University of Calgary Conjoint Health Research Ethics Board, REB18-0648) and all participants provided written consent for collection of urine and medical record data on mode of delivery and pregnancy outcomes. Participants were recruited at the Foothills Medical Centre (Calgary, AB) between May 2018 and July 2020. At recruitment, participants were classified based on gestational age [term (37-42 weeks) or preterm (<37 weeks)] and presence or absence of labour. Once delivered, participants were categorized into the following groups: term non-labour (TNL), n=32; term labour (TL), n=49, preterm non-labour controls (PTNL), n=15; threatened preterm labour with preterm delivery (TPTL-PTD), n=43, or threatened preterm labour with term delivery (TPTL-TD), n=44 (Fig 1).

**Fig 1.**
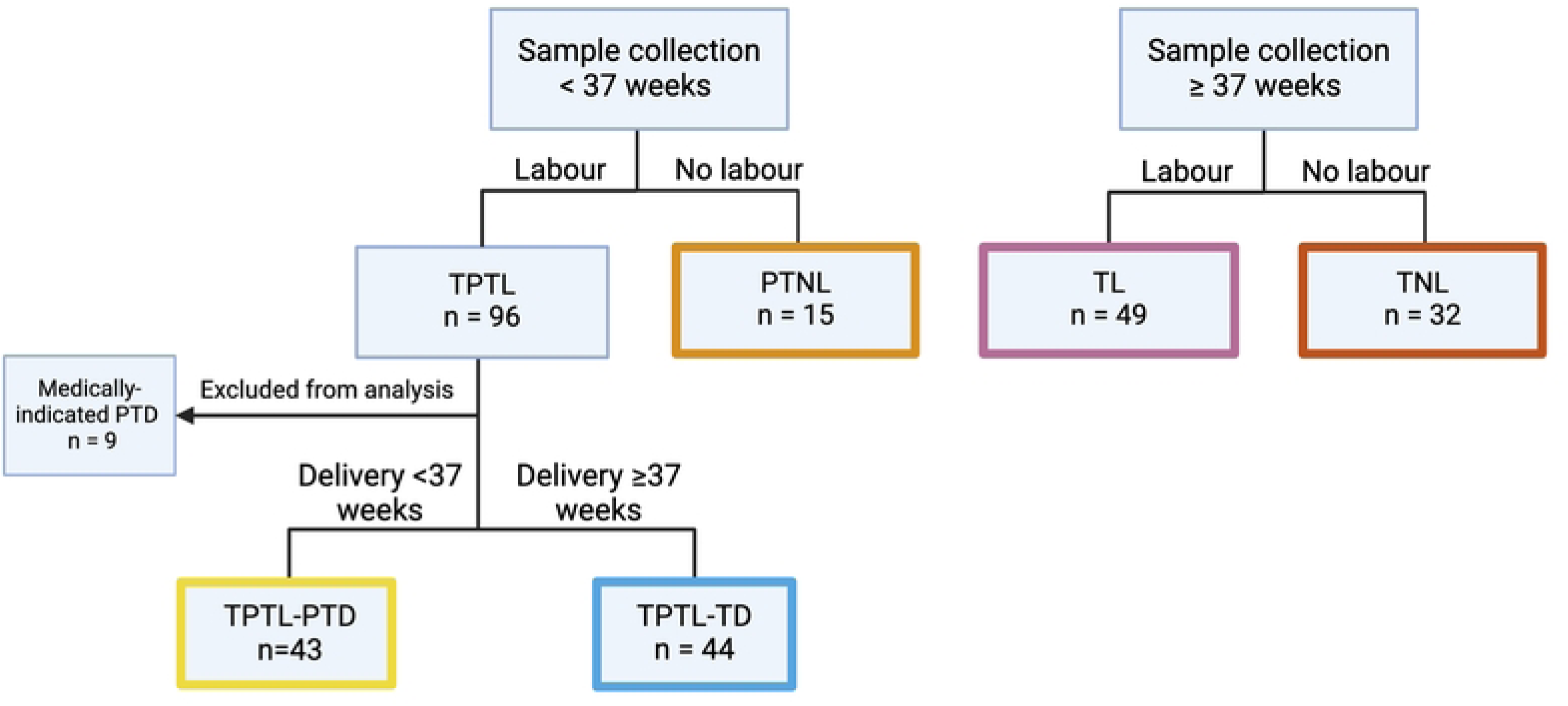
Participant grouping. Based on gestational age at sample collection (term vs preterm), presence or absence of labour, and outcome (preterm vs term delivery). TPTL = threatened preterm labour, PTNL = preterm non-labour controls, PTD = preterm delivery, TD = term delivery, TL = term labour, TNL = term non-labour. Created in BioRender. Wood, E. (2024) https://BioRender.com/i35i656

Term non-labour participants were recruited at presentation for induction or caesarean section at ≥37 weeks gestation with no symptoms of labour (no uterine contractions). Term labour was defined as spontaneous onset of labour, characterized by uterine contractions and cervical dilation, leading to delivery between 37-42 weeks gestation. Preterm non-labour controls were recruited either at presentation for non-delivery related reasons (n=12/15) or at presentation for induction or caesarean section at <37 weeks with no symptoms of labour (intact membranes and no reported contractions) (n=3/15). Diagnosis of threatened preterm labour was made based on clinical assessment by the attending physician and included symptoms such as spontaneous onset of uterine contractions or cramping, cervical changes including shortening and/or dilation, and/or premature rupture of membranes, prior to 37 weeks gestation. Participants with threatened preterm labour followed by a medically indicated preterm delivery, defined as delivery following induction of labour or caesarean section for fetal or maternal indications not related to labour, were excluded from the analysis. Deliveries were medically indicated for the following reasons: fetal heart rate abnormality, urinary retention, uterine dehiscence, and maternal medical conditions unrelated to pregnancy. An obstetrician blinded to the assay results was consulted to review cases where the outcome was unclear. Urine samples were collected in sterile containers at the time of recruitment and frozen at −80°C prior to analysis. Clinical information including cervical dilation and length, duration of ruptured membranes, and duration of contractions were recorded on study datasheets at the time of sample collection. Cervical score was calculated using the equation:

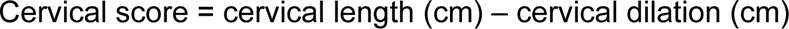

and used as a measure of cervical ripening and proximity to spontaneous delivery, as it has been demonstrated to perform similarly to the Bishop Score in predicting spontaneous delivery [23]. A cervical score of −10 indicates a fully dilated, fully effaced cervix, while a cervical score of 4 indicates a long and closed (i.e., unfavourable) cervix.

### Enzyme-linked immunosorbent assay (ELISA)

Urinary concentrations of 13,14-dihydro-15-keto PGF_2α_ (prostaglandin F metabolite; PGFM), tetranor-PGDM (t-PGDM), prostaglandin I metabolites (PGIM; 2,3-dinor-6-keto Prostaglandin F_1α_ and 20-carboxy-2,3-dinor-6-keto Prostaglandin F_1α_), and 8-isoprostane were measured by ELISA kit (Cayman Chemical, Ann Arbor, MI, item numbers #516671, #501001, #501100, and #516351, respectively). Prostaglandin E2 metabolite (PGEM) was measured using a PGE Metabolite ELISA kit (Cayman Chemical, Ann Arbor, MI, item number #514531) that converts the major circulating and urinary metabolites of PGE2, 13,14-dihydro-15-keto PGA_2_ and 13,14-dihydro-15-keto PGE_2_, to a single stable derivative that can be quantified by ELISA. Samples were run in duplicate, at dilutions of between 1:2 – 1:50 for PGFM, 1:5 – 1:50 for t-PGDM, 1:50 for PGIM, 1:25-1:50 for 8-isoprostane and 1:50 – 1:100 for PGEM. All samples were measured in duplicate. Assays were performed according to manufacturer’s protocol. Samples were assayed in random order and blinded to pregnancy outcome. ELISA data for PGFM, PGEM, t-PDGM, 8-isoprostane, PGIM and PGF_2α_ is provided in S1 Appendix.

### Urinary concentration correction

Urine concentration was normalized to specific gravity using the following formula [24]:

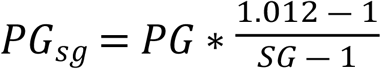

Where PG_sg_ is the specific gravity corrected prostaglandin metabolite concentration, 1.012 is the median specific gravity among all samples analyzed, PG is the measured prostaglandin concentration in pg/mL, and SG is the specific gravity of the sample. Specific gravity was measured using a Laxco™ Handheld Analog Clinical Refractometer (Thermo Fisher Scientific, Ottawa, ON, catalogue no. IRC200ATC) according to the manufacturer’s instructions.

Creatinine levels were measured for urine concentration normalization, however, due to a weak correlation between creatinine and gestational age (Spearman’s rank correlation: r(181)=0.31, p<0.0001; S2 Appendix), all subsequent analysis was conducted on specific gravity-corrected values.

### Reverse Phase Ultra High-Performance Liquid Chromatography-Mass Spectrometry

Lipid analysis was performed at the University of California San Diego (UCSD) Lipidomics Core using a mass spectrometry-based comprehensive eicosanoid panel. The eicosanoid panel consists of 147 lipids including, but not limited to, all major prostaglandins and their metabolites and three fatty acids (arachidonic acid, AA; eicosapentaenoic acid, EPA; docosahexaenoic acid, DHA). Full details of the eicosanoids measured in the panel are provided in S3 Appendix. Analysis was performed as described by Quehenberger and colleagues [25]. Briefly, eicosanoids were isolated from 400μL urine supplemented with 100μL internal standard mix by solid phase extraction using strata-x polymeric reverse phase columns (8B-S100-UBJ Phenomenex). Samples were then evaporated and reconstituted in 50μL of buffer (63% H2O, 37% acetonitrile, 0.02% acetic acid) and separated by reverse phase ultra-high performance liquid chromatography on an ACQUITY UPLC System Waters BEH-Shield column (2.1 x 100mm, 1.7μM; Waters, Milford, MA, USA). Eicosanoids were analyzed using a triple quadrupole linear ion trap mass spectrometer (Sciex 6500 Qtrap). Eicosanoids were quantitated by comparison with standards. Data analysis was performed using Analyst and MultiQuant software (Applied Biosystems). Mass spectrometry data is provided in S1 Appendix.

### Statistics and data presentation

Distributions of variables were assessed for normality using the Shapiro-Wilk test. Prostaglandin levels were corrected for urinary concentration by specific gravity and log2-transformed for normality. Differences in prostaglandin levels between groups were assessed by t-test. To assess the association between prostaglandin levels and cervical ripening the TL and TNL groups were combined and the PTNL, TPTL-PTD, and TPTL-TD groups were combined. Associations between prostaglandin levels and cervical score were assessed by Spearman’s correlation. Associations between prostaglandin levels and days from collection to delivery were assessed by linear regression. Data are presented as mean ± standard deviation (sd) for data normally distributed data and median ± interquartile range (IQR) for non-parametric data. Differences were considered significant with a 2-tailed p-value<0.05. Statistical analysis of demographic information was performed using R version 4.1.3. All other statistical analysis and visualization was performed using GraphPad Prism 9 software.

## Results

### Participant demographics

Demographic and clinical characteristics of participants are shown in Table 1. Compared to the TNL group, participants in the TL group had later median gestational age at collection (39.7 vs 38.6 weeks; p<0.001) and later median gestational age at delivery (39.7 vs 38.6 weeks; p<0.001). The mean maternal age of participants in the PTNL (mean = 31 ± 5.24 years; p = 0.030), TPTL-PTD (mean = 31.7 ± 4.88 years; p = 0.022), and TPTL-TD (mean = 31.3 ± 5.39 years; p = 0.009) groups, was lower than the TNL group (mean age = 34.5 ± 5.47 years).

**Table 1.** Demographic and clinical characteristics of participants.

|  | PTNL<br>(n=15) | TNL (n=32) | TL (n=49) | TPTL-PTD<br>(n=43) | TPTL-TD<br>(n=44) | p-value |
| --- | --- | --- | --- | --- | --- | --- |
| **Gestational age at collection (days)** | 223.0<br>(194.5-247.0) | 270.0<br>(267.5-273.0) | 278.0<br>(273.0-282.0) | 211.0<br>(182.0-247.0) | 220.0<br>(196.0-235.2) | TNL vs TL: $<0.001$<br>PTNL vs TPTL-PTD vs<br>TPTL-TD: $>0.05$ |
| Median<br>(IQR) |  |  |  |  |  |  |
| **Gestational age at delivery (days)**<br>Median (IQR) | 262 (256.5-273) | 270.0 (268.8-273.0) | 278.0 (274.0-282.0) | 219.0 (187.5-254.0) | 272.0 (266.7-275.0) | TNL vs TL: <0.001<br>TNL vs PTNL: >0.05<br>TNL vs TPTL-TD: >0.05<br>TL vs PTNL: <0.001<br>TL vs TPTL-TD: <0.001<br>PTNL vs TPTL-TD: 0.027 |
| **Time from collection to delivery (days)**<br>Median (IQR) | 33.5 (1-64.2) | 0 (0-1) | 0 (0-0) | 1 (0-5.5) | 55 (30.8-78.5) | NA |
| **Maternal age**<br>Mean ± sd | 31 ± 5.24 | 34.5 ± 5.47 | 32.3 ± 4.98 | 31.7 ± 4.88 | 31.3 ± 5.39 | TNL vs PTNL: 0.030<br>TNL vs TPTL-PTD: 0.022<br>TNL vs TPTL-TD: 0.009 |

Differences in maternal age between groups was assessed by ANOVA followed by pairwise t-test. Differences in all other variables were assessed by Wilcoxon rank sum test (no corrections for multiple testing). TNL = term non-labour, TL = term labour, PTNL = preterm non-labour controls, TPTL-PTD = threatened preterm labour – preterm delivery, TPTL-TD = threatened preterm labour – term delivery.

### Urinary prostaglandin levels with labour and gestation

The TL group was associated with significantly higher levels of PGFM, PGEM and PGF_2α_ compared to the TNL group (Table 2 and Fig 2). The levels of PGIM were significantly higher in TPTL-TD compared to the TPTL-PTD group. No other prostaglandin levels were different between TPTL-TD and TPTL-PTD groups and no gestational age differences in prostaglandin levels were observed (Fig 2).

**Fig 2.**
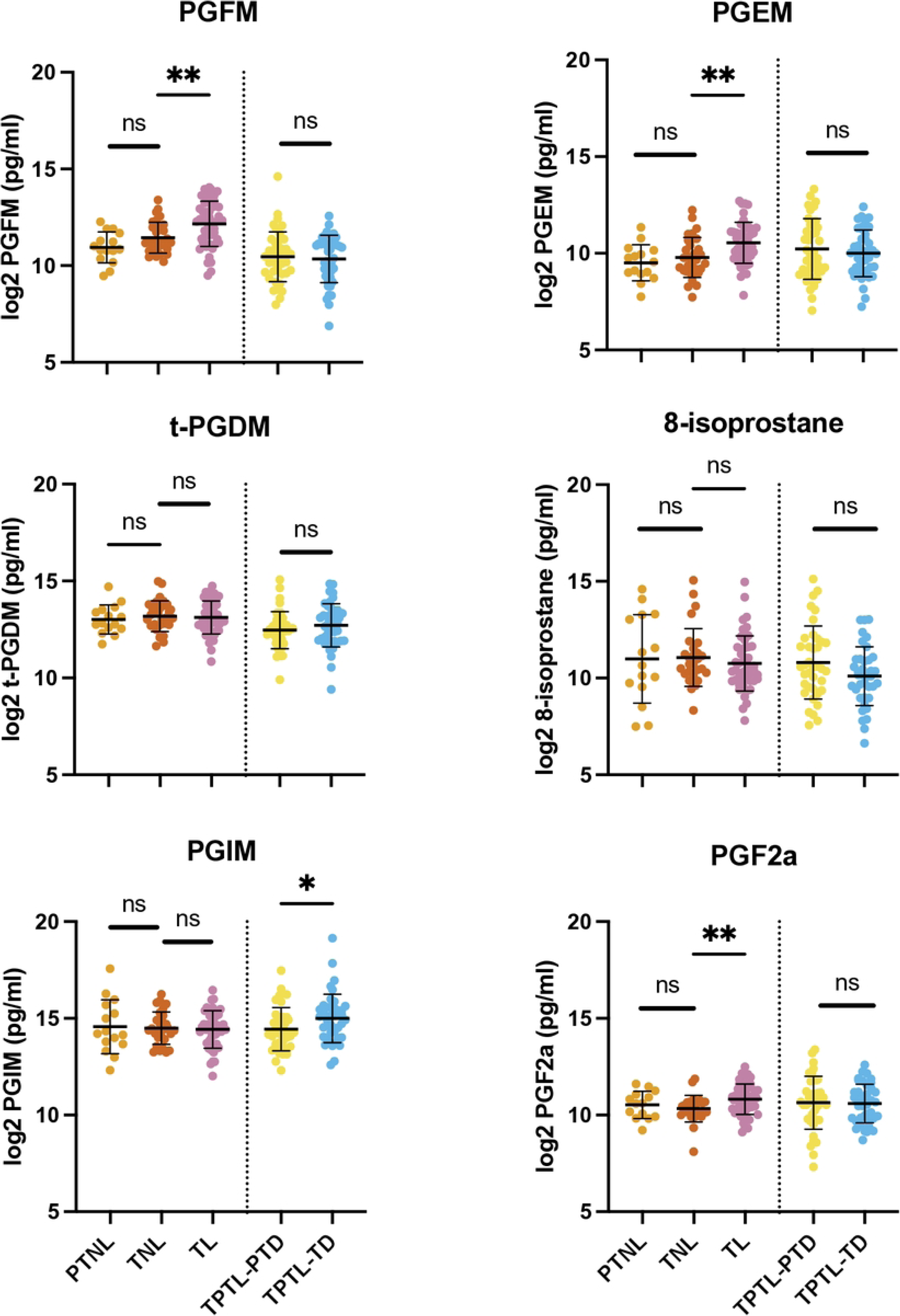
Prostaglandin and metabolite levels in maternal urine with and without labour. Prostaglandin levels were log2 transformed prior to statistical analysis. Differences analyzed by t-test. TNL = term non-labour, TL = term labour, PTNL = preterm non-labour controls, TPTL-TD = threatened preterm labour–term delivery, TPTL-PTD = threatened preterm labour-preterm delivery. *p<0.05, **p<0.01, ns = non-significant.

**Table 2.** Specific gravity corrected and log base 2 transformed urinary prostaglandin levels by group.

|  | TNL (n=32) | TL (n=49) | PTNL (n=15) | TPTL-PTD (n=43) | TPTL-TD (n=44) | p-value |
| --- | --- | --- | --- | --- | --- | --- |
| **PGFM** |  |  |  |  |  | TL vs TNL:<br>0.003<br>TPTL-PTD vs<br>TPTL-TD:<br>>0.05<br>PTNL vs TNL:<br>0.023 |
| Mean ± sd | 11.4 ± 0.799 | 12.2 ± 1.17 | 10.9 ± 0.805 | 10.5 ± 1.29 | 10.3 ± 1.23 |  |
| Total missing (%) | 0 (0%) | 0 (0%) | 0 (0%) | 1 (2.3%) | 0 (0%) |  |
| **PGEM** |  |  |  |  |  | TL vs TNL:<br>0.001<br>TPTL-PTD vs<br>TPTL-TD:<br>>0.05<br>PTNL vs TNL:<br>>0.05 |
| Mean ± sd | 9.79 ± 1.03 | 10.5 ± 1.06 | 9.51 ± 0.933 | 10.2 ± 1.57 | 10.0 ± 1.21 |  |
| Total missing (%) | 0 (0%) | 1 (2.0%) | 0 (0%) | 0 (0%) | 1 (2.3%) |  |
| **t-PGDM** |  |  |  |  |  | All p>0.05 |
| Mean ± sd | 13.2 ± 0.790 | 13.1 ± 0.846 | 13.0 ± 0.755 | 12.5 ± 0.955 | 12.7 ± 1.11 |  |
| Total missing (%) | 0 (0%) | 0 (0%) | 0 (0%) | 0 (0%) | 0 (0%) |  |
| **8-isoprostane** |  |  |  |  |  | All p>0.05 |
| Mean ± sd | 11.1 ± 1.52 | 10.8 ± 1.42 | 11.0 ± 2.29 | 10.8 ± 1.88 | 10.1 ± 1.52 |  |
| Total missing (%) | 4 (12.5%) | 2 (4.1%) | 0 (0%) | 5 (11.6%) | 5 (11.4%) |  |
| **PGIM** |  |  |  |  |  | TL vs TNL:<br>>0.05 |
| Mean ± sd | 14.5 ± 0.849 | 14.4 ± 0.965 | 14.6 ± 1.39 | 14.4 ± 1.12 | 15.0 ± 1.25 |  |
| Total missing (%) | 3 (9.4%) | 2 (4.1%) | 0 (0%) | 2 (4.7%) | 2 (4.5%) | TPTL-PTD vs TPTL-TD: 0.036<br>PTNL vs TNL: >0.05 |
| **PGF<sub>2α</sub>** |  |  |  |  |  | TL vs TNL: 0.009<br>TPTL-PTD vs TPTL-TD: >0.05<br>PTNL vs TNL: >0.05 |
| Mean ± sd | 10.3 ± 0.703 | 10.8 ± 0.786 | 10.5 ± 0.709 | 10.6 ± 1.37 | 10.6 ± 1.00 |  |
| Total missing (%) | 5 (15.6%) | 2 (4.1%) | 1 (6.7%) | 2 (4.7%) | 5 (11.4%) |  |

TNL = term non-labour, TL = term labour, PTNL = preterm non-labour controls, TPTL-PTD = threatened preterm labour – preterm delivery, TPTL-TD = threatened preterm labour – term delivery. Differences analyzed by t-test.

### Urinary prostaglandin levels and cervical ripening

At term, PGEM levels were positively associated with cervical ripening (R^2^ = 0.069, β = −0.099, p = 0.033; Fig 3a), while levels of PGFM, t-PGDM, 8-isoprostane, PGIM, and PGF_2α_ showed no changes (S4 Appendix, Supplemental Fig 2a-e). At preterm, t-PGDM and PGIM levels were negatively associated with cervical ripening (t-PGDM: R^2^= 0.081, β = 0.11 p = 0.017; PGIM: R^2^= 0.070, β = 0.12, p = 0.035; Fig 3b, c), while PGFM, PGEM, 8-isoprostane, and PGF_2α_ showed no association (S4 Appendix, Supplemental Fig 2f-I).

**Fig 3.**
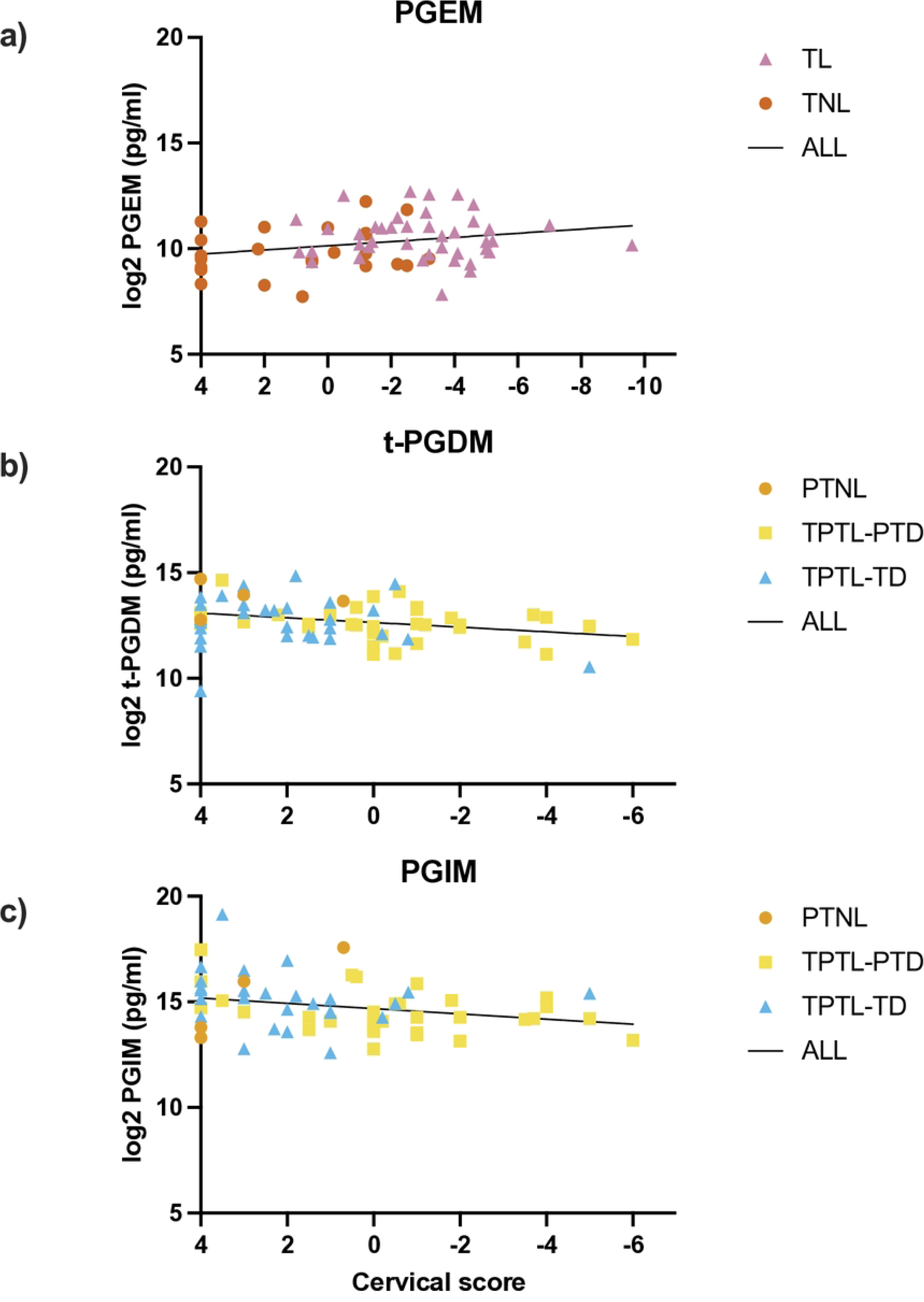
Association between urinary prostaglandin levels and cervical score. Analyzed by linear regression. a) PGEM: R^2^ = 0.069, β = −0.099, p = 0.034; b) t-PGDM: R^2^ = 0.081, β = 0.11, p = 0.017; c) PGIM: R^2^ = 0.070, β = 0.12, p = 0.035. TNL = term non-labour, TL = term labour, PTNL = preterm non-labour controls, TPTL-TD = threatened preterm labour–term delivery, TPTL-PTD = threatened preterm labour-preterm delivery. Regression equations represent all samples grouped together.

### Prostaglandin levels and time to delivery

In linear regression models of the associations between urinary prostaglandin levels and the number of days between sample collection and delivery in participants with threatened PTL, PGFM (R^2^ = 0.046, β = −0.008, p = 0.048), 8-isoprostane (R^2^ = 0.074, β = −0.015, p = 0.017), and PGF_2α_ (R^2^ = 0.065, β = −0.010, p = 0.022) levels were significantly associated with decreasing time to delivery, and PGIM levels were associated with increasing time to delivery (R^2^ = 0.062, β = 0.010, p =0.024; Fig 4).

**Fig 4.**
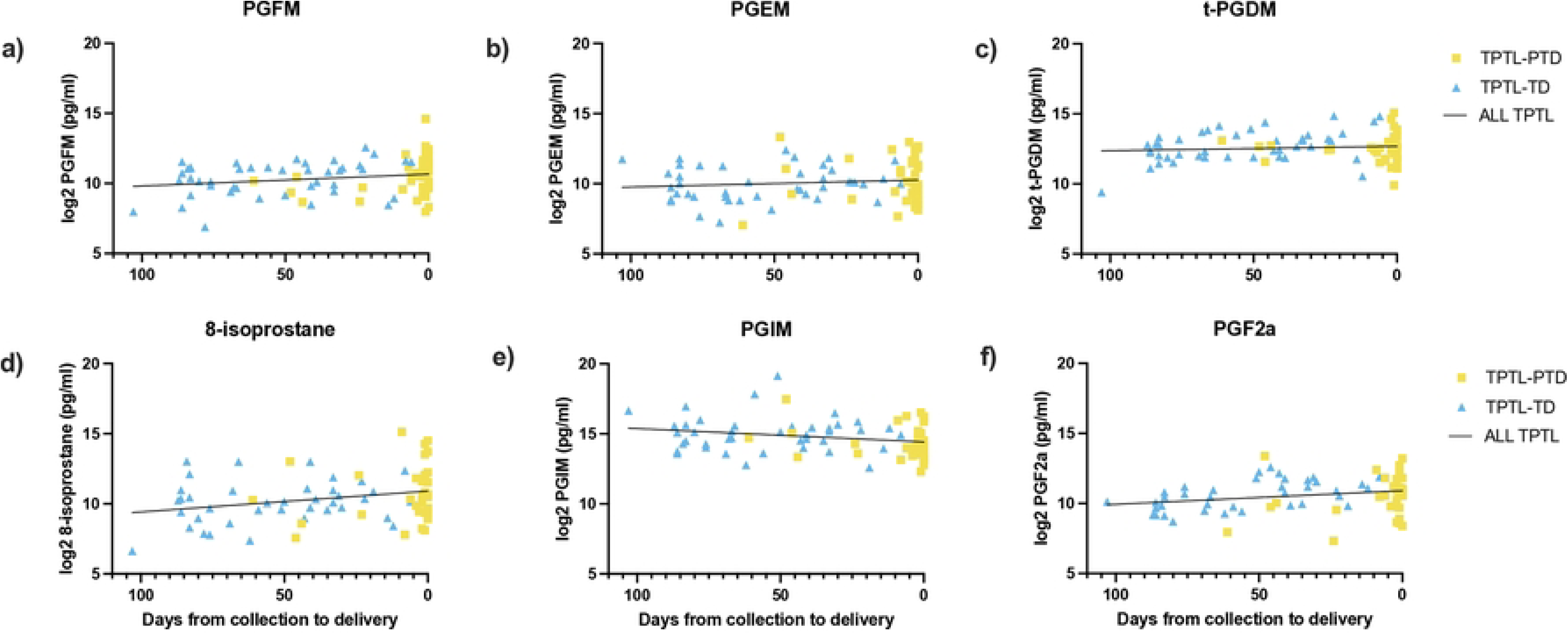
Association between urinary prostaglandin levels and timing to delivery in individuals with threatened preterm labour. Analyzed by linear regression. a) PGFM: R^2^ = 0.046, β = −0.008, p = 0.048; b) PGEM: R^2^ = 0.013, β = −0.005, p = 0.30; c) t-PGDM: R^2^ = 0.009, β = −0.003, p = 0.37; d) 8-isoprostane: R^2^ = 0.074, β = −0.015, p = 0.017; e) PGIM: R^2^ = 0.062, β = 0.010, p =0.024; f) PGF2a: R^2^ = 0.065, β = −0.010, p = 0.022. PTNL = preterm non-labour controls, TPTL-TD = threatened preterm labour–term delivery, TPTL-PTD = threatened preterm labour-preterm delivery

### Discovery cohort eicosanoid profile

To determine additional eicosanoids that may be present in maternal urine, a subset of n=24 urine samples were selected for a discovery analysis using a mass-spectrometry-based eicosanoid panel. Groups were composed of n=6 threatened preterm labour with preterm delivery (TPTL-PTD), n=6 preterm non-labour controls (PTNL), n=6 term labour (TL), and n=6 term non-labour (TNL) (Table 3). Of the 147 eicosanoids measured, 66 were not detected in any sample. A total of 81/147 eicosanoids and fatty acids were detected in at least one urine sample and ranged from concentrations of 0.01 pmol/mL to 1016.17 pmol/mL. Of the 81 eicosanoids, 4 were detected in all samples (9,10-diHOME, 9,10-EpOME, 9-HODE, and 13-HODE) and 20 were detected in at least 50% of samples. Eicosanoids detected in at least 50% of samples include members of the prostaglandin, hydroxyeicosatetraenoic acid (HETE), epoxyeicosatrienoic acid (EET), dihydroxy-octadecenoic acid (DiHOME), dihydroxy-eicosatrienoic acid (diHETrE), isoprostane, and nitro fatty acid eicosanoid families.

**Table 3.** Discovery cohort demographics.

|  | PTNL (n=6) | TPTL-PTD (n=6) | TNL (n=6) | TL (n=6) | p-value |
| --- | --- | --- | --- | --- | --- |
| **Gestational age at collection (weeks)** | 30.1 (26-34.4) | 29 (26.4-33.5) | 39.3 (38.6-40.4) | 39.1 (39-39.6) | TNL vs TL >0.05<br>PTNL vs TPTL-PTD >0.05 |
| **Gestational age at delivery (weeks)** | 39.1 (38.6-40.5) | 29.2 (26.6-33.5) | 39.3 (38.6-40.4) | 39.1 (39-39.6) | TNL vs TL >0.05<br>TNL vs PTNL >0.05<br>TL vs PTNL >0.05 |
| **Time from collection to delivery (days)** | 58 (38.8-93) | 1.5 (0.25-2) | 0 (0-0.75) | 0 (0-0) | TNL vs TL >0.05 |
| **Maternal age** | 32 (27.5-35) | 30 (22-35.8) | 36 (30.5-39.2) | 31.5 (28.8-33.5) | All >0.05 |

Data represented as median (IQR). TNL = term non-labour, TL = term labour, PTNL = preterm non-labour control, TPTL-PTD = threatened preterm labour – preterm delivery, NR = not reported, NA = not applicable.

## Discussion

### Summary

Here we have shown that metabolites of prostaglandins F_2α_ and E_2_, PGFM and PGEM, are increased in maternal urine with term labour compared to term non-labour. However, as evidenced by the lack of gestational age-related changes in prostaglandin metabolite levels, none of the prostaglandins measured here appear to increase leading up to labour onset. Additionally, we show that PGIM was the only metabolite that displayed a significant, albeit small, difference in individuals with threatened PTL who delivered preterm compared to those who delivered at term. The evidence provided here therefore suggests that none of the prostaglandin biomarkers measured are likely to be *clinically* useful for prediction of labour onset, either at term or in the instance of threatened preterm labour.

### PGFM and PGEM levels are higher with term labour, but do not change prior to labour

That prostaglandins PGF_2α_ and PGE_2_ and their metabolites increase during labour is well established in the literature and has been consistently demonstrated in various biofluids including amniotic fluid [26–38], maternal blood [32, 39–47], and maternal urine [32, 48–50]. That we were not able to find any changes in prostaglandin levels with gestational age (preterm non-labour vs term non-labour), is also generally consistent with previous studies that have measured prostaglandin metabolites in maternal blood [32, 42, 51–53]. Several studies conducted using amniotic fluid samples, however, have reported increases in PGF_2α_ and PGE_2_ around term, prior to labour onset [27,28, 35, 36, 54, 55], suggesting that biomarkers predictive of labour onset may be present in this biofluid. Although maternal urine is appealing as a minimally invasive biomarker option, it may not provide the best reflection of the physiological processes occurring prior to labour. It may be possible that uterine prostaglandin production does increase prior to labour onset, and that urinary output of prostaglandin metabolites simply does not reflect this increase. Although this seems unlikely to be the case, as previous studies have demonstrated that increases in prostaglandin metabolites show up rapidly (i.e., at least within 2 hours) in urine following systemic increases in these prostaglandins [56,57]. An alternative explanation is that PGF_2α_ and PGE_2_ metabolites do increase with term labour, but do not change prior to labour onset, which could support the premise that prostaglandin metabolite production is a consequence of labour onset, rather than a trigger for this process, as suggested by others [58].

### Prostaglandins and cervical ripening

We also noted PGEM levels were associated with cervical ripening at term, but not preterm. There is extensive literature detailing the association between exogenous administration of PGE_2_ and cervical ripening [59–61], however few others have investigated the association between endogenous levels of PGE_2_ or its metabolites, and degree of cervical ripening [62]. The results demonstrated here provide support for a role for PGE_2_ in the physiological process of cervical ripening, although the question of whether the increase in PGE_2_ metabolites is a cause or a consequence of cervical ripening remains.

### Prostaglandin metabolite levels may not be useful as predictors of preterm delivery in individuals with threatened PTL

Of special interest in this study was whether prostaglandin metabolite levels could be used to differentiate between threatened preterm labour that resolves and eventually ends in a term delivery from threatened preterm labour that results in a preterm delivery. Previous studies have identified higher levels of PGE_2_ metabolites and PGFM in amniotic fluid [63] and PGFM in maternal plasma [64] in individuals with PTL leading to preterm delivery compared to those who eventually delivered at term. However, others have found no difference in plasma PGFM levels between these two groups [52]. We did not find any differences in urinary PGE_2_ or PGF_2α_ metabolite levels between those with threatened PTL and preterm delivery compared to those with threatened PTL and term delivery, however, we did find higher levels of prostaglandin I_2_ metabolites (PGIM) in TPTL-TD compared to TPTL-PTD. Prostaglandin I_2_ (PGI_2_) is known to induce smooth muscle relaxation in the uterus [11,12], and has been implicated in pregnancy maintenance [65]. It is therefore possible that lower levels of PGI_2_ could contribute to labour onset via loss of quiescent signaling in the uterus, although we did not see any changes in PGIM prior to labour onset when comparing PTNL to TNL. In our analysis of the association between PGIM and timing to delivery in individuals with threatened PTL, PGIM accounted for a small percentage (R^2^ = 0.062) of the variance in time to delivery, making this metabolite unlikely to be useful as a biomarker for preterm delivery in individuals with threatened PTL.

### Role for diverse eicosanoids in labour

Beyond prostaglandins there is limited information on the levels and trajectories of eicosanoids throughout pregnancy and a poor understanding of how these molecules may contribute to the process of labour onset. In the present investigation, we have identified several non-prostaglandin eicosanoids present in maternal urine at levels comparable to, or higher than, maternal urinary prostaglandin levels, some of which could potentially be useful as biomarkers for labour. Among the eicosanoids identified, include members of the lipoxygenase (LOX)-derived HETEs and CYP450-derived EETs and diHOMEs. Notably, previous investigations have reported that higher plasma levels of the LOX-derived eicosanoid, 5-HETE, in the second trimester are associated with increased risk of spontaneous preterm birth [66,67]. Aung and colleagues also reported associations between plasma levels of 12,13-dihydroxy-octadecenoic acid (12,13-diHOME) and 9,10-dihydroxy-octadecenoic acid (9,10-diHOME) and increased risk of preterm birth and found that LOX and CYP450 pathway eicosanoids demonstrated better predictive capability for spontaneous preterm birth than COX pathway eicosanoids (i.e., prostaglandins) [66]. Further, Borkowski et al. (2020) report an association between spontaneous preterm birth and LOX pathway metabolites in obese pregnant individuals, but not in “normal weight” pregnancies, suggesting that this association may be BMI-dependent [68]. Further investigation into the role of these eicosanoids in pregnancy and labour, and other factors that may influence their levels, could potentially uncover better biomarkers for conditions like preterm birth.

### Limitations

In this analysis, we included participants with preterm labour and participants with preterm premature rupture of membranes (PPROM) in one group under threatened PTL. Although both preterm labour and PPROM are events that contribute to spontaneous delivery, it is possible that the molecular pathways leading to the onset of each of these events is distinct. In the future, studies with larger numbers should aim to analyse these groups separately.

## Conclusion

The study herein suggests that while changes in prostaglandins (PGF_2α_) and prostaglandin metabolites (PGFM, PGEM, PGIM) are associated with human labour, we did not identify any biomarkers that increased prior to labour onset and thus would have utility as predictors for term or preterm labour. However, we have identified an additional 20 eicosanoid molecules present in at least half of our discovery cohort samples, that may play a role in pregnancy and labour. We have provided here the mass spectrometry lipidomics data and suggest that future studies should further investigate the role of these novel non-prostaglandin eicosanoids in pregnancy and labour.

## Data Availability

All relevant data are within the manuscript and its Supporting Information files.

## Acknowledgements

We thank our study participants and study support staff, in particular Jill Putnam. We would also like to thank Emma Walsh, whom we remember fondly.

## Supporting Information

**S1 Appendix. ELISA and mass spectrometry data.**

**S2 Appendix. Supplemental Fig 1. Urinary creatinine levels throughout pregnancy by gestational age at sample collection.**

**S3 Appendix. UCSD Lipidomics Core eicosanoid panel.**

**S4 Appendix. Supplemental Fig 2. Urinary prostaglandin metabolite levels not associated with cervical score in term or preterm pregnancy.**

## References

1. Aronsson A, Ulfgren A-K, Ståbi B, Stavreus-Evers A, Gemzell-Danielsson K. The effect of orally and vaginally administered misoprostol on inflammatory mediators and cervical ripening during early pregnancy. Contraception. 2005;72(1):33–9.

2. Osmers R, Rath W, Adelmann-Grill BC, Fittkow C, Szeverényi M, Kuhn W. Collagenase activity in the human cervix uteri after prostaglandin E2 application during the first trimester. Eur J Obstet Gynecol Reprod Biol. 1991;42(1):29–32.

3. Yoshida M, Sagawa N, Itoh H, Yura S, Takemura M, Wada Y, et al. Prostaglandin F2α, cytokines and cyclic mechanical stretch augment matrix metalloproteinase-1 secretion from cultured human uterine cervical fibroblast cells. Molecular Human Reproduction. 2002;8(7):681–7.

4. Senior J, Marshall K, Sangha R, Clayton JK. In vitro characterization of prostanoid receptors on human myometrium at term pregnancy. Br J Pharmacol. 1993;108(2):501–6.

5. Wikland M, Lindblom B, Wilhelmsson L, Wiqvist N. Oxytocin, prostaglandins, and contractility of the human uterus at term pregnancy. Acta Obstet Gynecol Scand. 1982;61(5):467–72.

6. Li W, Unlugedik E, Bocking AD, Challis JRG. The Role of Prostaglandins in the Mechanism of Lipopolysaccharide-Induced proMMP9 Secretion from Human Placenta and Fetal Membrane Cells. Biology of Reproduction. 2007;76(4):654–9.

7. McLaren J, Taylor DJ, Bell SC. Prostaglandin E2-dependent production of latent matrix metalloproteinase-9 in cultures of human fetal membranes. Molecular Human Reproduction. 2000;6(11):1033–40.

8. Ulug U, Goldman S, Ben-Shlomo I, Shalev E. Matrix metalloproteinase (MMP)-2 and MMP-9 and their inhibitor, TIMP-1, in human term decidua and fetal membranes: the effect of prostaglandin F2α and indomethacin. Molecular Human Reproduction. 2001;7(12):1187-93.

9. Karim SM, Trussell RR, Patel RC, Hillier K. Response of pregnant human uterus to prostaglandin-F2-alpha-induction of labour. Br Med J. 1968;4(5631):621-3.

10. Fetalvero KM, Zhang P, Shyu M, Young BT, Hwa J, Young RC, et al. Prostacyclin primes pregnant human myometrium for an enhanced contractile response in parturition. The Journal of Clinical Investigation. 2008;118(12):3966–79.

11. Omini C, Pasargiklian R, Folco GC, Fano M, Berti F. Pharmacological activity of PGI2 and its metabolite 6-oxo-PGF1alpha on human uterus and fallopian tubes. Prostaglandins. 1978;15(6):1045–54. Wilhelmsson, L., Wikland, M., & Wiqvist, N. (1981). PGH2, TxA2 and PGI2 have potent and differentiated actions on human uterine contractility. Prostaglandins, 21(2), 277–286.

12. Wilhelmsson L, Wikland M, Wiqvist N. PGH2, TxA2 and PGI2 have potent and differentiated actions on human uterine contractility. Prostaglandins. 1981;21(2):277–86.

13. Bennett PR, Elder MG, Myatt L. The effects of lipoxygenase metabolites of arachidonic acid on human myometrial contractility. Prostaglandins. 1987;33(6):837–44.

14. Eghtedari AR, Safizadeh B, Vaezi MA, Kalantari S, Tavakoli-Yaraki M. Functional and pathological role of 15-Lipoxygenase and its metabolites in pregnancy and pregnancy-associated complications. Prostaglandins & Other Lipid Mediators. 2022;161:106648.

15. Kikut J, Komorniak N, Ziętek M, Palma J, Szczuko M. Inflammation with the participation of arachidonic (AA) and linoleic acid (LA) derivatives (HETEs and HODEs) is necessary in the course of a normal reproductive cycle and pregnancy. J Reprod Immunol. 2020;141:103177.

16. Moore TA, Ahmad IM, Zimmerman MC. Oxidative Stress and Preterm Birth: An Integrative Review. Biological Research For Nursing. 2018;20(5):497–512.

17. Howson CP, Kinney MV, Lawn J. Born too soon: the global action report on preterm birth. 2012.

18. Hornaday KK, Wood EM, Slater DM. Is there a maternal blood biomarker that can predict spontaneous preterm birth prior to labour onset? A systematic review. PLoS One. 2022;17(4):e0265853.

19. Menon R, Torloni MR, Voltolini C, Torricelli M, Merialdi M, Betrán AP, et al. Biomarkers of Spontaneous Preterm Birth: An Overview of The Literature in the Last Four Decades. Reproductive Sciences. 2011;18(11):1046–70.

20. Gazmararian JA, Petersen R, Jamieson DJ, Schild L, Adams MM, Deshpande AD, et al. Hospitalizations during pregnancy among managed care enrollees. Obstet Gynecol. 2002;100(1):94–100.

21. Scott CL, Chavez GF, Atrash HK, Taylor DJ, Shah RS, Rowley D. Hospitalizations for severe complications of pregnancy, 1987-1992. Obstet Gynecol. 1997;90(2):225–9.

22. Wood EM, Hornaday KK, Slater DM. Prostaglandins in biofluids in pregnancy and labour: A systematic review. PLoS One. 2021;16(11):e0260115.

23. Newman RB, Goldenberg RL, Iams JD, Meis PJ, Mercer BM, Moawad AH, et al. Preterm prediction study: comparison of the cervical score and Bishop score for prediction of spontaneous preterm delivery. Obstet Gynecol. 2008;112(3):508–15.

24. Boeniger, M. F., Lowry, L. K., & Rosenberg, J. (1993). Interpretation of urine results used to assess chemical exposure with emphasis on creatinine adjustments: a review. Am Ind Hyg Assoc J, 54(10), 615–627. doi:10.1080/15298669391355134

25. Quehenberger, O., Armando, A. M., Brown, A. H., Milne, S. B., Myers, D. S., Merrill, A. H., … Dennis, E. A. (2010). Lipidomics reveals a remarkable diversity of lipids in human plasma. J Lipid Res, 51(11), 3299–3305. doi:10.1194/jlr.M009449

26. Maddipati KR, Romero R, Chaiworapongsa T, Zhou SL, Xu Z, Tarca AL, et al. Eicosanomic profiling reveals dominance of the epoxygenase pathway in human amniotic fluid at term in spontaneous labor. FASEB Journal. 2014;28(11):4835–46.

27. Salmon JA, Amy J-J. Levels of prostaglandin F2α in amniotic fluid during pregnancy and labour. Prostaglandins. 1973;4(4):523–33.

28. Hibbard BM, Sharma SC, Fitzpatrick RJ, Hamlett JD. Prostaglandin F2alpha concentrations in amniotic fluid in late pregnancy. The Journal of obstetrics and gynaecology of the British Commonwealth.

29. Hillier K, Calder AA, Embrey MP. Concentrations of prostaglandin F2alpha in amniotic fluid and plasma in spontaneous and induced labours. The Journal of obstetrics and gynaecology of the British Commonwealth. 1974;81(4):257–63.

30. Dray F, Frydman R. Primary prostaglandins in amniotic fluid in pregnancy and spontaneous labor. American journal of obstetrics and gynecology. 1976;126(1):13–9.

31. Keirse MJ, Mitchell MD, Turnbull AC. Changes in prostaglandin F and 13,14-dihydro-15-keto-prostaglandin F concentrations in amniotic fluid at the onset of and during labour. British journal of obstetrics and gynaecology. 1977;84(10):743–6.

32. Satoh K, Yasumizu T, Fukuoka H, Kinoshita K, Kaneko Y, Tsuchiya M, et al. Prostaglandin F2 alpha metabolite levels in plasma, amniotic fluid, and urine during pregnancy and labor. American journal of obstetrics and gynecology. 1979;133(8):886–90.

33. Romero R, Baumann P, Gomez R, Salafia C, Rittenhouse L, Barberio D, et al. The relationship between spontaneous rupture of membranes, labor, and microbial invasion of the amniotic cavity and amniotic fluid concentrations of prostaglandins and thromboxane B2 in term pregnancy. American journal of obstetrics and gynecology. 1993;168(6 Pt 1):1654–8.

34. Romero R, Baumann P, Gonzalez R, Gomez R, Rittenhouse L, Behnke E, et al. Amniotic fluid prostanoid concentrations increase early during the course of spontaneous labor at term. American Journal of Obstetrics and Gynecology. 1994;171(6):1613–20.

35. Romero R, Munoz H, Gomez R, Parra M, Polanco M, Valverde V, et al. Increase in prostaglandin bioavailability precedes the onset of human parturition. Prostaglandins Leukotrienes and Essential Fatty Acids. 1996;54(3):187–91.

36. Lee SE, Romero R, Park I-S, Seong HS, Park C-W, Yoon BH. Amniotic fluid prostaglandin concentrations increase before the onset of spontaneous labor at term. The journal of maternal-fetal & neonatal medicine : the official journal of the European Association of Perinatal Medicine, the Federation of Asia and Oceania Perinatal Societies, the International Society of Perinatal Obstetricians. 2008;21(2):89–94.

37. Peiris HN, Romero R, Vaswani K, Gomez-Lopez N, Tarca AL, Gudicha DW, et al. Prostaglandin and prostamide concentrations in amniotic fluid of women with spontaneous labor at term with and without clinical chorioamnionitis. Prostaglandins, Leukotrienes and Essential Fatty Acids. 2020;163:102195.

38. Takahashi N, Okuno T, Fujii H, Makino S, Takahashi M, Ohba M, et al. Up-regulation of cytosolic prostaglandin E synthase in fetal-membrane and amniotic prostaglandin E2 accumulation in labor. PloS one. 2021;16(4):e0250638-e.

39. Mitchell MD, Flint AP, Bibby J, Brunt J, Arnold JM, Anderson AB, et al. Plasma concentrations of prostaglandins during late human pregnancy: influence of normal and preterm labor. J Clin Endocrinol Metab. 1978;46(6):947–51.

40. Mitchell MD, Ebenhack K, Kraemer DL, Cox K, Cutrer S, Strickland DM. A sensitive radioimmunoassay for 11-deoxy-13, 14-dihydro-15-keto-11, 16-cyclo-prostaglandin E2: application as an index of prostaglandin E2 biosynthesis during human pregnancy and parturition. Prostaglandins Leukotrienes & Medicine. 1982;9(5):549–57.

41. Haning RV, Jr., Barrett DA, Alberino SP, Lynskey MT, Donabedian R, Speroff L. Interrelationships between maternal and cord prolactin, progesterone, estradiol, 13,14-dihydro-15-keto-prostaglandin F2alpha, and cord cortisol at delivery with respect to initiation of parturition. American Journal of Obstetrics & Gynecology. 1978;130(2):204–10.

42. Dubin NH, Johnson JW, Calhoun S, Ghodgaonkar RB, Beck JC. Plasma prostaglandin in pregnant women with term and preterm deliveries. Obstetrics & Gynecology. 1981;57(2):203–6.

43. Fuchs A-R, Husslein P, Sumulong L, Fuchs F. The origin of circulating 13,14-dihydro-15-keto-prostaglandin F2α during delivery. Prostaglandins. 1982;24(5):715–22.

44. Fuchs AR, Goeschen K, Husslein P, Rasmussen AB, Fuchs F. Oxytocin and the initiation of human parturition III. Plasma concentrations of oxytocin and 13,14-dihydro-15-keto-prostaglandin F2 alpha in spontaneous and oxytocin-induced labor at term. American Journal of Obstetrics and Gynecology. 1983;147(5):497–502.

45. Sellers SM, Mitchell MD, Anderson AB, Turnbull AC. The influence of spontaneous and induced labour on the rise in prostaglandins at amniotomy. Br J Obstet Gynaecol. 1984;91(9):849–52.

46. Nagata I, Sunaga H, Furuya K, Makimura N, Kato K. Changes in the plasma prostaglandin F2 alpha metabolite before and during spontaneous labor and labor induced by amniotomy, oxytocin and prostaglandin E2. Endocrinol Jpn. 1987;34(2):153–9.

47. Johnston TA, Greer IA, Kelly RW, Calder AA. Plasma prostaglandin metabolite concentrations in normal and dysfunctional labour. Br J Obstet Gynaecol. 1993;100(5):483–8.

48. Granstrom E, Kindahl H. Radioimmunoassay for urinary metabolites of prostaglandin F2alpha. Prostaglandins. 1976;12(5):759–83.

49. Hamberg M. Quantitative studies on prostaglandin synthesis in man III. Excretion of the major urinary metabolite of prostaglandins F1α and F2α during pregnancy. Life Sciences. 1974;14(2):247–52.

50. Ichikawa M, Minami M. Correlations between corticotropin-releasing hormone and L-3,4-dihydroxyphenylalanine in plasma, and dopamine and prostaglandins in urine in pregnant mothers. Biogenic Amines. 1999;15(2):287–306.

51. Ghodgaonkar RB, Dubin NH, Blake DA, King TM. 13, 14-dihydro-15-keto-prostaglandin F2alpha concentrations in human plasma and amniotic fluid. American Journal of Obstetrics & Gynecology. 1979;134(3):265–9.

52. Sellers SM, Mitchell MD, Bibby JG, Anderson AB, Turnbull AC. A comparison of plasma prostaglandin levels in term and preterm labour. Br J Obstet Gynaecol. 1981;88(4):362–6.

53. Yamaguchi M, Mori N. 6-Keto prostaglandin F1 alpha, thromboxane B2 and 13,14-dihydro-15-keto prostaglandin F levels in maternal and fetal plasma. Prostaglandins Leukotrienes & Medicine. 1984;15(3):325–36.

54. Johnson DA, Manning PA, Hennam JF, Newton JR, Collins WP. The concentration of prostaglandin f2alpha in maternal plasma, foetal plasma and amniotic fluid during pregnancy in women. Acta Endocrinologica. 1975;79(3):589–97.

55. Nieder J, Augustin W. Increase of prostaglandin E and F equivalents in amniotic fluid during late pregnancy and rapid PG F elevation after cervical dilatation. Prostaglandins Leukotrienes and Medicine. 1983;12(3):289–97.

56. Hamberg M, Samuelsson B. On the metabolism of prostaglandins E 1 and E 2 in man. J Biol Chem. 1971;246(22):6713–21.

57. Okayasu I, Kuroiwa H, Shinkawa K, Hayashi K, Sato S, Iwata N, et al. Significant increase in prostaglandin E-major urinary metabolite with physical exercise suggesting muscle inflammation. All Life. 2023;16(1):2167868.

58. MacDonald PC, Casey ML. The accumulation of prostaglandins (PG) in amniotic fluid is an aftereffect of labor and not indicative of a role for PGE2 or PGF2 alpha in the initiation of human parturition. The Journal of clinical endocrinology and metabolism. 1993;76(5):1332–9.

59. Calder AA. Prostaglandins and Biological Control of Cervical Function. Australian and New Zealand Journal of Obstetrics and Gynaecology. 1994;34(3):347–51.

60. Crane JMG, Bennett KA. A Meta-analysis of Controlled-Release Prostaglandin for Cervical Ripening and Labour Induction. Journal SOGC. 2000;22(9):692–8.

61. Orr L, Reisinger-Kindle K, Roy A, Levine L, Connolly K, Visintainer P, et al. Combination of Foley and prostaglandins versus Foley and oxytocin for cervical ripening: a network meta-analysis. American Journal of Obstetrics and Gynecology. 2020;223(5):743.e1-.e17.

62. Keirse MJ, Turnbull AC. E prostaglandins in amniotic fluid during late pregnancy and labour. The Journal of obstetrics and gynaecology of the British Commonwealth. 1973;80(11):970–3.

63. Romero R, Wu YK, Sirtori M, Oyarzun E, Mazor M, Hobbins JC, et al. Amniotic fluid concentrations of prostaglandin F2 alpha, 13,14-dihydro-15-keto-prostaglandin F2 alpha (PGFM) and 11-deoxy-13,14-dihydro-15-keto-11, 16-cyclo-prostaglandin E2 (PGEM-LL) in preterm labor. Prostaglandins. 1989;37(1):149–61.

64. Weitz CM, Ghodgaonkar RB, Dubin NH, Niebyl JR. Prostaglandin F metabolite concentration as a prognostic factor in preterm labor. Obstetrics and Gynecology. 1986;67(4):496–9.

65. Korita D, Sagawa N, Itoh H, Yura S, Yoshida M, Kakui K, et al. Cyclic Mechanical Stretch Augments Prostacyclin Production in Cultured Human Uterine Myometrial Cells from Pregnant Women: Possible Involvement of Up-Regulation of Prostacyclin Synthase Expression. The Journal of Clinical Endocrinology & Metabolism. 2002;87(11):5209–19.

66. Aung MT, Yu Y, Ferguson KK, Cantonwine DE, Zeng L, McElrath TF, et al. Prediction and associations of preterm birth and its subtypes with eicosanoid enzymatic pathways and inflammatory markers. Scientific Reports. 2019;9(1):17049.

67. Ramsden CE, Makrides M, Yuan ZX, Horowitz MS, Zamora D, Yelland LN, et al. Plasma oxylipins and unesterified precursor fatty acids are altered by DHA supplementation in pregnancy: Can they help predict risk of preterm birth? Prostaglandins Leukot Essent Fatty Acids. 2020;153:102041.

68. Borkowski K, Newman JW, Aghaeepour N, Mayo JA, Blazenović I, Fiehn O, et al. Mid-gestation serum lipidomic profile associations with spontaneous preterm birth are influenced by body mass index. PLOS ONE. 2020;15(11):e0239115.

